# iSupport-J, the Japanese version of an internet-based self-learning and psychological assessment program for dementia caregivers: study protocol for a randomized, waiting-list controlled study

**DOI:** 10.1101/2022.11.16.22282333

**Authors:** Shingo Yamashita, Yuma Yokoi, Makoto Matsui, Kazumi Nozaki, Chinatsu Fujimaki, Ayumu Wada, Naoya Iwata, Norio Sugawara, Yoshie Omachi

## Abstract

**Background:** The number of people with dementia in Japan will estimate to increase to roughly 20% of those aged 65 and over (approximately 7 million people) by 2025. Around the world, the number of people with dementia is increasing by 7.7 million per year, and caregivers’ depression, stress, social isolation, and sleep disorders due to the burden of caregiving are also increasing. Economic losses worldwide due to physical and mental health problems of dementia caregivers, reduced work hours due to caregiving, and job loss are estimated to be $252 billion in 2010. In addition, the risk of abuse against the caregiver increases as the caregiver is affected by physical and mental illness. Psychosocial interventions such as cognitive behavioral therapy have reportedly reduced caregivers’ mental distress and improved their health. Since caregivers have significant time and physical limitations due to caregiving, it is promising that interventions using the internet, which have few limitations in terms of location and time, will be helpful, along with its low cost.

**Methods:** This is a two-arm, randomized, waitlist-controlled trial. Participants will be randomized with a 1:1 ratio to either the immediate or late access group. The early intervention group will be able to use iSupport for three months, followed by allocation and follow-ups until six months. In the waitlist group, iSupport can be available for three months from the end of the Month 3 evaluation. Scheduled evaluation periods are Months 1, 3, and 6.

**Discussion:** We plan to begin recruiting for the trial in January 2021. We plan to finish the inclusion by June 2021 and complete all data collection by December 2021. Once all data collection is complete, we plan to analyze the data by April 2022; we aim to publish the results in a manuscript by the end of 2022.

**Trial Registration:** UMIN-CTR, UMIN000042957, registered on January 9, 2021.

## Background

The elderly population is growing worldwide, and the number of people aged 65 and over will likely reach 36.7 million by 2025 [1]. Also, in Japan, one of the most aged countries, the number of patients with dementia will reportedly increase to 20% of those aged 65 and over (7 million people) by 2025 [2]. Globally, the number of patients with dementia worldwide was estimated to be 46.8 million in 2015 [3]. Furthermore, the number will assumably double every 20 years, rising to 74.7 million by 2030 and 131.5 million by 2050[3].

Family members and other informal caregivers are primarily responsible for caring for people with dementia, especially in developing countries or countries characterized by familial practices. In Japan, informal care accounts for 70% of the total care provided, including 54% provided by family members living with the family, partly due to familial practices[4].

Caring for a person with dementia requires much care, from money management to bathing and diaper changing, which are both mentally and physically taxing for the caregivers. Therefore, they are at high risk of developing mental disorders such as depression, anxiety, and various physical diseases [5–7]. Furthermore, depression and anxiety of caregivers are associated with abuse against caregivers [8], and reducing depression and anxiety of caregivers is considered helpful for both patients with dementia and caregivers. Psychosocial interventions to help caregivers cope with challenging behaviors and stressful situations, communicate with people with dementia, and improve their ability to seek support from others may be effective in improving caregivers’ mental health and well-being [6,9-10]. Likewise, educating caregivers about the symptoms of dementia, how it progresses, how to control the symptoms, and how to use social resources is believed to reduce depressive symptoms and the sense of the burden of caregivers, and education for caregivers may be positive in maintaining the mental health of caregivers [11]. Additionally, psychological interventions such as cognitive-behavioral therapy for caregivers have been reported to reduce the mental distress of caregivers, such as anxiety and depression, and improve their health status.

Caregivers have significant time and physical constraints due to caregiving, interventions using the internet, which has fewer location and time constraints, are expected to be beneficial and inexpensive. In addition, the number of caregivers who have access to face-to-face interventions is limited due to reluctance and prejudice against asking non-family members to take care of family members.

For the abovementioned reasons, internet interventions are considered reasonable and supportive for caregivers in burden. In Japan, many people, including the elderly, use the internet, and we can intervene with caregivers via the internet, even if they are aged spouses.

A meta-analysis of RCTs(Randomized controlled trials) of Internet-based support interventions for dementia caregivers found that the interventions predominantly improved depressive symptoms and perceived stress and improved anxiety and caregiver self-efficacy. Alternatively, there were no significant improvements in the caregiver burden, coping ability, caregiver response to challenging behavior, or quality of life [12–15]. The World Health Organization(WHO) recognizes dementia as a public health priority[16]. In May 2017, the World Health Assembly endorsed the Global action plan on the public health response to dementia 2017– 2025 [17]. Support for dementia caregivers is one of the main goals of the action plan.

Therefore WHO has developed iSupport, an online knowledge/skills training program aimed at improving the caregiver’s mental health and coping skills, with the help of international experts. iSupport expectedly improves caregivers’ mental health and coping skills through relaxation skills, cognitive restructuring, and problem-solving thinking using cognitive behavioral therapy techniques.

In Japan, the Comprehensive Strategy for Dementia Policy Promotion (the Orange Plan) has been in place since 2012 as a policy for the aging society. However, it was revised in 2017 (the New Orange Plan)[18] to create a society where the elderly with dementia can live independently. The plan calls for promoting efforts to reduce the mental and physical burdens of family members and other caregivers, R&D of nursing care models, and disseminating the results.

In the New Orange Plan, caregiver support is one of the main pillars of promoting dementia-friendly communities, and we decided to introduce iSupport in Japan. In doing so, we followed the WHO adaptation guide and translated it into Japanese according to the Japanese cultural background. With the cooperation of the Association of People with Dementia and Their Families, a dementia family association in Japan, and nursing care specialists, an open-label, uncontrolled study was conducted to evaluate the Japanese version of the iSupport (iSupport-J) and the Psychological Evaluation and Questionnaire (electronic patient-reported outcome; ePRO) system. We surveyed the ease of understanding and usage of iSupport-J and the ePRO system and made changes to the presentation and system.

In this paper, we describe the design of an RCT to assess the effectiveness of iSupport-J, the Japanese version of iSupport.

## Methods

### Study Design

This study is a two-arm, randomized, waitlist-controlled trial.

The first thing after enrollment is to randomize eligible family caregivers to Group A (initial intervention group) or Group B(waitlist control group). Group A will be able to access iSupport for three months, followed by the randomization immediately. On the other hand, Group B will be able to access iSupport after three months waitlist term (see Figure 1). Participants in the low-burden group with low CES-D or ZBI scores during the screening are not eligible for the study but can still use iSupport as Group C (reference group). The study will follow the Consolidation Standards of Reporting Trials (CONSORT) guidelines [19].

**Figure.**
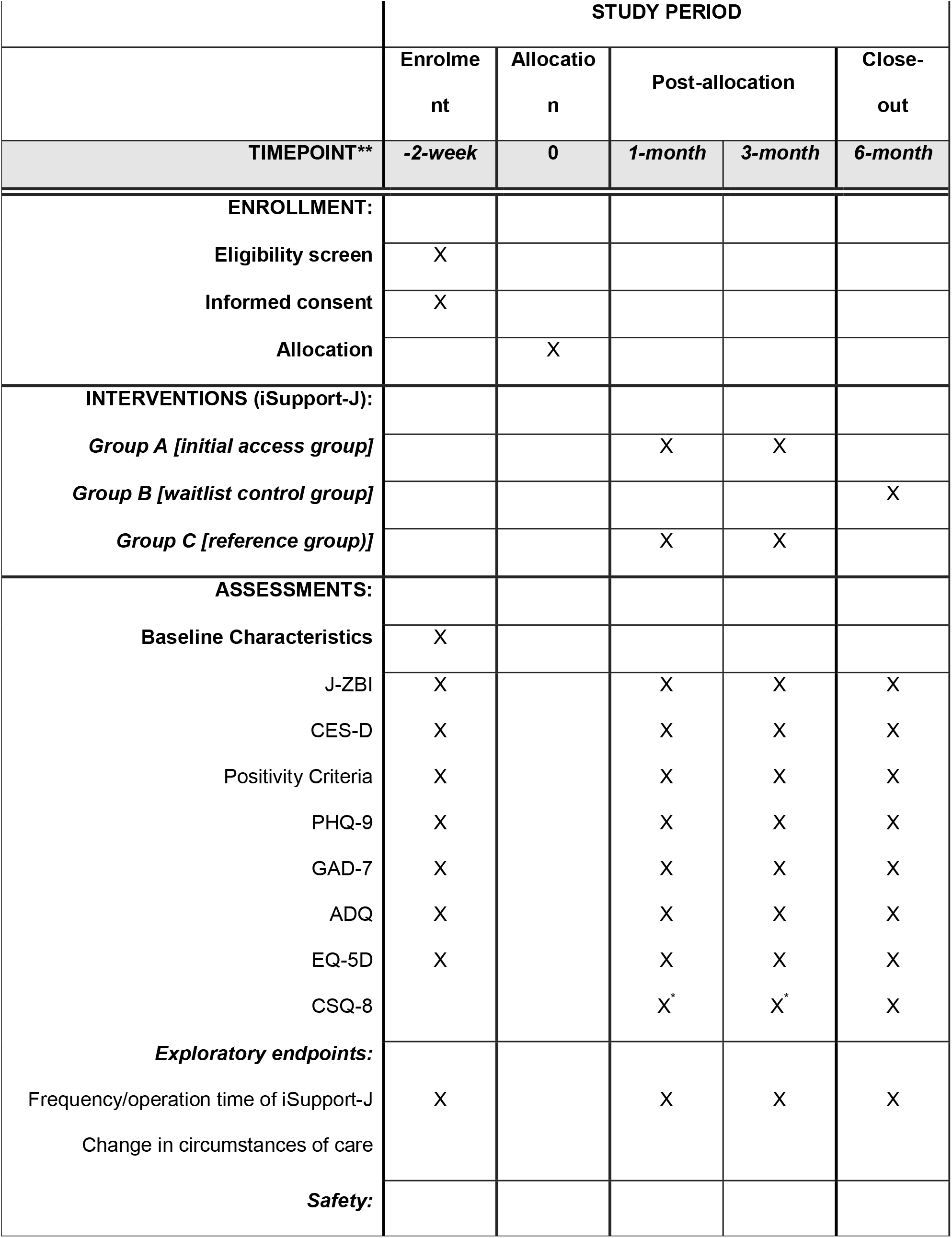
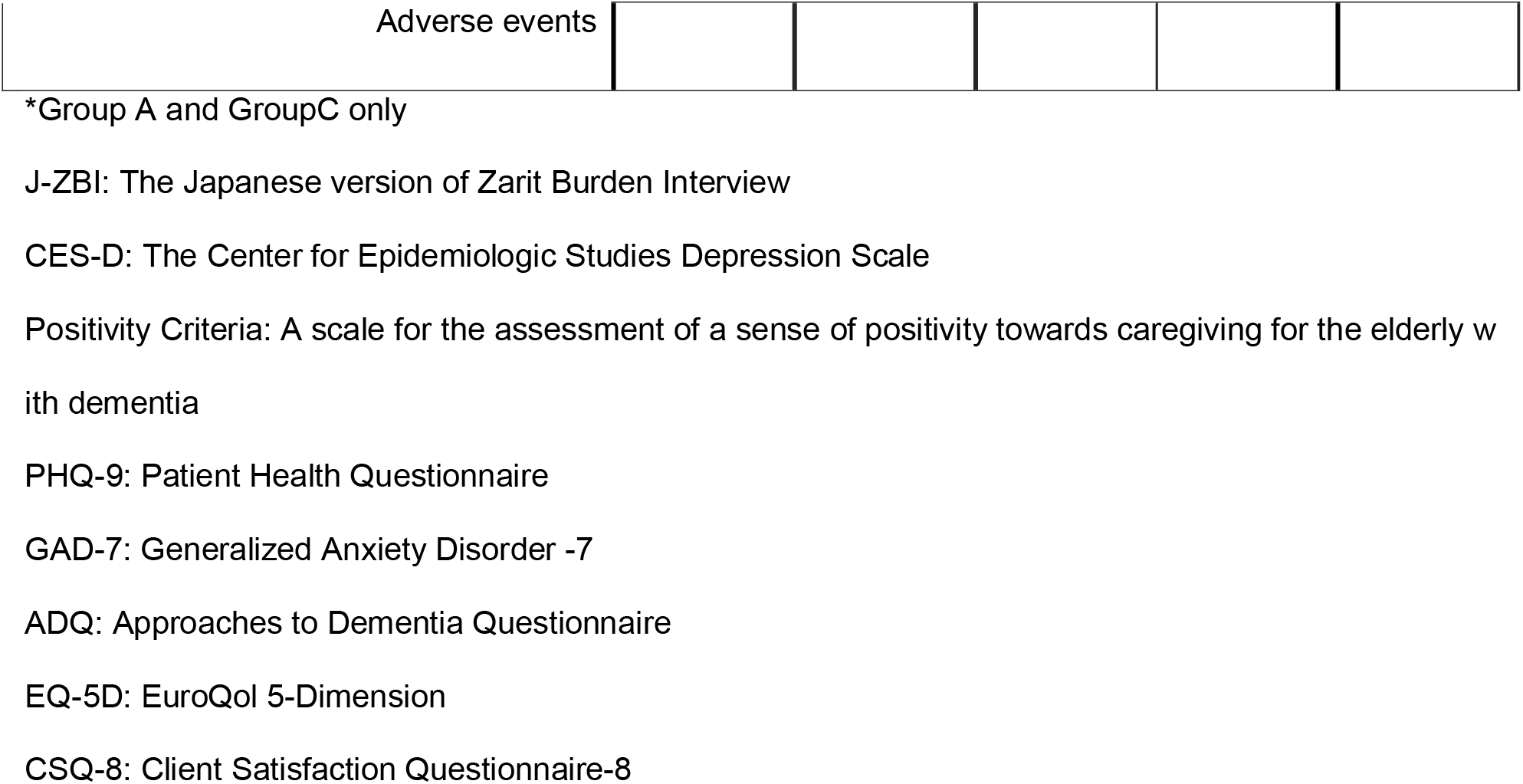
Schedule of iSupport intervention study.

### Participants

Participants are screened and included if they (1) are aged 18 years and older, (2) are self-reported caregiver whose care recipient has been diagnosed with dementia, (3) are a person who has access to the internet, (4) score 21 or more on the Japanese version of the Zarit Burden Interview (J-ZBI) [20–21], (5) score four or more the Japanese version of the Center for Epidemiological Studies Depression (CES-D) scale[22-23]. Participants are excluded from the study if they (1) score 26 or more on the CES-D scale, (2) score 15 or more on the Japanese version of the Generalized Anxiety Disorder-7 (GAD-7) scale [24–25], and (3) are judged inappropriate by the investigators. Besides, applicants with J-ZBI score of 20 or less and CES-D score of 3 or less will not be included in the RCT but will be evaluated as a reference group (low-burden group).

### Randomization

As soon as the iSupport-J office staff receives participants’ inputs in the ePRO, randomization starts automatically using a computer, according to stratified allocation with two covariates: gender (male/female) and caregiver/recipient relationship (spouse/other).

Participants assigned to Groups A and C will begin the iSupport intervention immediately, while those assigned to Group B will receive the iSupport intervention after a 3-month waiting period.

## iSupport-J

iSupport is based on the WHO’s Mental Health Gap Action Programme’s evidence-based guidelines for dementia caregivers. The contents comprise the following five chapters and a summary.

Module 1. Introduction to Dementia

Module 2. Being a Caregiver

Module 3. Caring for me

Module 4. Providing everyday care

Module 5. Dealing with Challenging Behaviors

Before the start of this trial, we created a Japanese version of iSupport (iSupport-J) per the guidelines of WHO. We checked the understandability of iSupport-J and the ease of use of ePRO through a pilot study (N=11). We made modifications to make it easier for the elderly and adapt it to Japanese culture and system. In addition, we also added audio and video guidance for relaxation and mindfulness meditation, as well as various pictures for proactive involvement.

User data stored in the personal information management system automatically links to the ePRO system. The system is usable with a web browser and is guaranteed to work on all standard terminals (listed in “Supported OS and Browsers”).

The participants by themselves will register to participate in the study and register their personal information. The iSupport-J Office will access the personal information registered by the participants and contact them as necessary.

Participants log in with an ePRO ID and password and answer psychological evaluations and questionnaires online or study via the iSupport-J. The iSupport-J Office will contact the participants according to the participants’ status of log-ins, response to psychological evaluations and questionnaires, and achievements in the iSupport-J.

## Interventions

Caregivers who wish to participate in the study will apply via the iSupport-J website. After registration in the iSupport-J system, the participants will answer the questionnaire with the ePRO system on the internet. The researcher will send the informed consent forms, explain the study over the phone if necessary, and obtain written consent. Those who meet the exclusion criteria will be dropped out and will get notified.

In case of participants with suspected severe depression (CES-D scale score of 26 or more) or anxiety (GAD-7 scale score of 15 or more), the iSupport-J Office recommends they see psychiatrists or get local mental health and welfare services.

Participants who meet the inclusion criteria and are eligible for random allocation will divide into Group A (initial intervention group) and Group B (waitlist control group).

Group A will engage in the iSupport-J for three months, and participants report psychological evaluations and questionnaires via the ePRO system at baseline (at the beginning of the intervention), one month, three months (at the end of the intervention), and six months (post-intervention period). Group B will wait for three months, and then they can use iSupport-J for three months. Items and timing of evaluations are as same as Group B, at baseline, one month, three months (at the beginning of the intervention), and six months (at the end).

Participants who are ineligible for allocation but not excluded will be evaluated as Group C (reference group) according to the same schedule as Group A. However, the data in Group C will not be used for the primary analysis but for exploratory analyses.

During the iSupport-J intervention period, participants will self-learn at their convenience. However, if two or three weeks pass without logging in, the iSupport-J system will send a reminder email to the participants. When three weeks have passed without logging in to e-learning, the iSupport-J system will also contact the iSupport-J Office, and the iSupport-J Office will remind the participants by phone or email.

When it is time to conduct the psychological evaluation and questionnaire, the iSupport-J system will notify the participants. The allowance of the input period will be two weeks. Suppose the input by the participants has yet to be confirmed even after three days remain until the due date. In that case, the iSupport-J system will contact the iSupport-J Office, and the iSupport-J Office will remind the subject by phone or email.

## Outcome measures

The Japanese version of the Zarit Burden Interview (J-ZBI: a scale that measures physical and psychological burdens and economic difficulties as caregiver burdens) is the 21-item version of the Dementia Caregiver Affirmation Scale (a scale for assessing a sense of positivity toward caregiving for the elderly with dementia)[26]. J-ZBI is the primary endpoint of this study. We also adapted the Japanese version of the Center for Epidemiological Studies Depression scale (CES-D Scale, a scale to measure the severity of depressive symptoms) and the Japanese version of the Generalized Anxiety Disorder-7 scale(GAD-7 scale, a scale to measure anxiety) for evaluating caregivers’ depression, affirmation, and anxiety.

Other endpoints will be caregivers’ awareness of person-centered care, quality of life (QOL), and satisfaction with the application. We used each of the following scales: the Japanese version of the Approaches to Dementia Questionnaire [27-28](ADQ; evaluation of caregivers’ mind and attitude of person-centered care), the Japanese version of the EuroQol 5-Dimension [29-30](EQ-5D; QOL rating scale), and the Client Satisfaction Questionnaire [31-32](CSQ-8J; a questionnaire for evaluating application satisfaction). In addition, evaluations include a questionnaire to survey changes in care status and use of social resources because of using the iSupport-J system and time spent using the iSupport-J system. We will scrutinize the program’s implementation rate, completion rate, and usage time through the iSupport-J system.

## Sample size calculation

Assuming an alpha of 0.05, a statistical power (1-beta) of 0.80 in a two-tailed test, and a standardized effect size of 0.33, considered the clinically meaningful difference in reducing the J-ZBI for the target population, 80 subjects per group are the estimated minimum sample size. We planned to recruit 104 subjects per group, considering possible missing data and dropouts.

## Covariates

Covariates regarding caregivers are age, gender, working status, number of hours per week for providing care, number of family caregivers, and the relationship between the caregiver and the cared-for person. Covariates about the cared-for person are the diagnosis of dementia, other medical histories, former hospitalization due to dementia, number of years since dementia, level of care required, level of support, and level of social resources they use. Although it may not be any covariate, we collect data about iSupport-J itself by following questions; “Where did you learn about iSupport-J?”, “What are the good and bad points of iSupport-J?” and “how much do you learn by using iSupport-J or other media?”.

## Statistical analysis

Distribution of demographic variables (e.g., gender, age, relationship to the caregiver, duration of care) and reference values (J-ZBI, CES-D, GAD-7, ADQ, EQ-5D, CSQ-8J, CGS) of the subject population participating in this study will be summarized. Descriptive statistics will identify the attributes and trends of participants. We will calculate frequencies and proportions for categorical variables, and for continuous-type variables, we will compute summary statistics for each intervention group (mean ± standard deviation, median, minimum and maximum).

The analysis will be based on the intention-to-treat principle for whomever was randomized into either Group A or B. We will conduct a mixed model for repeated measures (MMRM) analysis to detect changes in J-ZBI, the 21-item version of the Dementia Caregiver Affirmation Scale, CES-D, GAD-7, values from baseline to the Month 3 visit. The MMRM model includes the fixed effects of group, time point (Months 1, 3, 6), group-by-time interaction, and baseline characteristics (gender, caregiver/recipient relationship). The primaly analysis per protocol set will be a sensitivity analysis and all analyses of secondary outcomes. All analyses will be performed using SAS V.9.4.

## Interim analysis

We plan an interim analyses of potential sample size increase and safety when the 80th participant finishes the Month 3 assessment. In principle, we will not suspend enrollment during the interim analysis unless the pace of enrollment is extremely higher than expected at the time of the interim analysis. Whether or not to conduct an unscheduled enrollment pause will be decided after discussion by the study team and the principal investigator.

The allocator will blind the allocation group to 80 cases in each group for interim analysis (labeling the groups in a manner that is not analogous to the group name, e.g., R group and L group) and provide the statistician with data from baseline to Month 3.

The statistician will evaluate the mean values of the background information at baseline for each group or the entire group and present the results to the principal investigator.

The statistician will also evaluate the mean values for each group or the entire group for information after the baseline that does not directly affect efficacy (continuation rate) and present the results to the principal investigator.

The same analysis as the primary analysis will be performed for efficacy, using the J-ZBI score at repeated time points (baseline, Month 1, and Month 3), group (early intervention group and waiting list group), age, relationship to the caregiver, self-reported hours of care per week, level of care required by the caregiver, and caregiver’s mental health status. The MMRM estimation method uses the restricted maximum likelihood method (REML). In addition, the Kenward-Roger method is used to estimate the degrees of freedom. To identify the variance-covariance structure, we first estimate in Unstructured (UN). If there is no convergence, we estimate in Heterogeneous CS (CSH) and Compound Symmetry (CS) sequentially. However, no statistical tests are performed.

## Discussion

Recognizing that supporting family and other informal caregivers is an important priority, the World Health Organization (WHO) created iSupport. iSupport is originally in English but now has been translated and adapted to local cultures in various countries, and several RCTs are evaluating its effectiveness.

In Japan, the care system for patients with dementia was based on the municipality deciding the necessary services based on the user’s condition until 2000 when the Long-Term Care Insurance Law was enacted. Users can now receive comprehensive healthcare and medical services from various entities. One of the objectives of iSupport is to enable the appropriate use of external resources. In Japan, there are relatively good one-stop services such as long-term care insurance, but the effectiveness of iSupport will be affected by whether it encourages appropriate use. We plan to investigate whether there are any changes in the usage of services before and after using iSupport.

Research on this iSupport worldwide starts to disseminate. The first RCT was conducted in India [33], followed by the Netherlands [34] and Portugal [35]. In India, the control group was education only; in the Netherlands, it was the waitlist group. Adaptations have also been made in Australia [36], Brazil [37], Switzerland [38] for future efficacy studies.

According to a report from India [39], of the 74 patients in the iSupport group, only 29 could complete the follow-up assessment after three months, and more than 60% dropped out.

Since we acknowledged a high probability of dropout if participants were allocated to the e-books and never able to use iSupport-J, we decided to conduct an RCT in Japan with an iSupport group and a waitlist group, as we did in the Netherlands.

In Japan, the system automatically send reminder emails at the end of 2 and 3 weeks to motivate people to engage in iSupport if they still need to log in to iSupport. Based on the results of this study in India, it was considered essential to work on lowering the dropout rate. There was a report that adding phone call contacts to email reminders and monetary incentives did increase follow-up rates[40], suggesting that direct phone calls are necessary in addition to email reminders. That reminder methods need to be modified slightly.

Finally, we would like to conclude by asserting that we have ensured the safety of our research. We decided to exclude participants who had depression or anxiety above a certain level. In such cases, the medical personnel would call them and introduce them to the family associations for dementia in Japan and recommend that they visit a medical institution.

## Conclusions

In this study, we plan to verify the effectiveness of iSupport. If this study confirms the effectiveness of iSupport, we plan to make it available to more people.

## Trial Status

This study is currently recruiting participants.

## Data Availability

All data produced in the present study are available upon reasonable request to the authors

https://isupport-j.org/

## List of Abbreviations

ADQ: Approaches to Dementia Questionnaire
CES-D: Center for Epidemiologic Study Depression Scale
CONSORT: Consolidation Standards of Reporting Trials
CSQ-8J: Client Satisfaction Questionnaire
EQ-5D: EuroQol 5-Dimension
GAD-7: Generalized Anxiety Disorder scale-7-item
GEE: Generalized estimating equations
RCT: Randomized controlled trial
T0: Pre measurement
T1: First follow-up (1month)
T2: Second follow-up (3months)
WHO: World Health Organization
ZBI: Zarit Burden Interview

## Ethics approval and consent to participate

The local institutional review board has approved approved our reseach (National Center of Neurology and Psychiatry Ethics Committee, A2020-030). The principal investigator, the research coordinator, or the research assistant will be responsible for conducting the informed consent process with the study participants. All subjects must give consent to participate in the trial. Any relevant changes in the study protocol and the informed consent will be sent to the Institutional Review Board as a protocol amendment. All the subjects’ identities will be protected with a unique code that only the principal investigator can access. Before starting the trial, the protocol was registered in UMIN-CTR (UMIN000042957).

## Consent for publication

Not applicable

## Availability of data and material

Not applicable.

## Author contributions

SY, YY, NS, and YO developed the design and methodology.

SY, YY, MM, KN, CF, AW, NI, NS, and YO created the Japanese version of iSupport. SY, YY, XX, NS, and YO developed the analysis plan.

All authors contributed to drafting the article and read and approved the final manuscript.

## Acknowledgments

The authors would like to thank WHO for permitting us to create the Japanese version of iSupport. The authors would also like to thank the expert panel, the Association of People with Dementia and their Families, and the Kodaira City government for cooperating in preparing the Japanese version of iSupport.

## References

1. National Institute of Population and Social Security Research: Report on Japan’s Future Population Projections. 2017. http://www.ipss.go.jp/ppzenkoku/j/zenkoku2017/pp29_ReportALL.pdf. Accessed February 3, 2021.

2. Cabinet Office, Government of Japan. White paper on aging society. 2017. https://www8.cao.go.jp/kourei/whitepaper/w-2017/zenbun/29pdf_index.html. (in Japanese). Accessed February 3, 2021.

3. Altzheimer’s Disease International : World Alzheimer Report. 2015. 10-45.

4. Ministry of Health, Labour and Welfare, Government of Japan. Overview of the 2019 National Survey on Living Standards. 2019 https://www.mhlw.go.jp/toukei/saikin/hw/k-tyosa/k-tyosa19/dl/05.pdf (in Japanese) Accessed February 3, 2021.

5. Kaddour L, Kishita N, Schaller A. A meta-analysis of low-intensity cognitive behavioral therapy-based interventions for dementia caregivers. Int Psychogeriatrics [Internet]. 2019 ;31(07):961–976. Available from: https://www.cambridge.org/core/product/identifier/S1041610218001436/type/journal_article

6. World Health Organization. Supporting Informal Caregivers of People Living With Dementia. 2015. https://www.who.int/mental_health/neurology/dementia/dementia_thematicbrief_informal_care.pdf Accessed February 3, 2021

7. Givens JL, Mezzacappa C, Heeren T, Yaffe K, Fredman L. Depressive symptoms among dementia caregivers: Role of mediating factors. Am J Geriatr Psychiatry. 2014;22(5):481–488.

8. Cooper C, Blanchard M, Selwood A, Walker Z, Livingston G. Family carers’ distress and abusive behaviour: Longitudinal study. Br J Psychiatry. 2010 Jun;196(6):480–485.

9. Ory MG, Hoffman RR 3rd, Yee JL, Tennstedt S, Schulz R. Prevalence and impact of caregiving: a detailed comparison between dementia and nondementia caregivers. Gerontologist. 1999 Apr;39(2):177–85.

10. Gilhooly KJ, Gilhooly MLM, Sullivan MP, McIntyre A, Wilson L, Harding E, et al. A meta-review of stress, coping and interventions in dementia and dementia caregiving. BMC Geriatr. 2016 May 18;16(1).

11. Terayama H, Sakurai H, Namioka N, Jaime R, Otakeguchi K, Fukasawa R, et al. Caregivers’ education decreases depression symptoms and burden in caregivers of patients with dementia. Psychogeriatrics [Internet]. 2018 Sep 1;18(5):327–33. Available from: https://doi.org/10.1111/psyg.12337

12. Leng M, Zhao Y, Xiao H, Li C, Wang Z. Internet-Based Supportive Interventions for Family Caregivers of People With Dementia: Systematic Review and Meta-Analysis. J Med Internet Res. 2020 Sep;22(9):e19468.

13. Boots LMM, de Vugt ME, van Knippenberg RJM, Kempen GIJM, Verhey FRJ. A systematic review of Internet-based supportive interventions for caregivers of patients with dementia. Int J Geriatr Psychiatry. 2014 Apr;29(4):331–44.

14. Parra-Vidales E, Soto-Pérez F, Perea-Bartolomé MV, Franco-Martín MA, Muñoz-Sánchez JL. Online interventions for caregivers of people with dementia: a systematic review. Actas Esp Psiquiatr. 2017 May;45(3):116–26.

15. Egan KJ, Pinto-Bruno ÁC, Bighelli I, Berg-Weger M, van Straten A, Albanese E, et al. Online Training and Support Programs Designed to Improve Mental Health and Reduce Burden Among Caregivers of People With Dementia: A Systematic Review. J Am Med Dir Assoc. 2018 Mar;19(3):200-206.e1.

16. Dementia: A Public Health Priority. World Health Organization; 2012. https://apps.who.int/iris/bitstream/handle/10665/75263/9789241564458_eng.pdf?sequence=1 Accessed February 3, 2021.

17. Global Action Plan on the Public Health Response to Dementia: 2017-2025. World Health Organization;2017. https://apps.who.int/iris/bitstream/handle/10665/259615/9789241513487-eng.pdf?sequence=1 Accessed February 3, 2021.

18. The Ministry of Health, Labor and Welfare, Government of Japan; 2017. The Comprehensive Strategy for Dementia Policy Promotion (New Orange Plan) https://www.mhlw.go.jp/file/06-Seisakujouhou-12300000-Roukenkyoku/nop101.pdf Accessed February 3, 2021.

19. Schulz K, Altman D, Moher D, CONSORT Group. CONSORT 2010 Statement: Updated guidelines for reporting parallel group randomised trials. BMC Med. 2010; 8:18 [doi: 10.1186/1741-7015-8-18] [Medline: 20334633]

20. Zarit S, Reever K, Bach-Peterson J. Relatives of the Impaired Elderly: Correlates of Feelings of Burden. Gerontologist. 1981; 20:649–55.

21. Arai, Y. The Japanese version of Zarit Burden Interview (J-ZBI) and its short version (J -ZBI_8). Nihon Rinsho. Japanese Journal of Clinical Medicine, 2004; 62(4), 45–50.

22. Radloff, L. S. The CES-D scale. Appl Psychol Meas, 1977; 1. 385–401.

23. Satoru, S. Tatsuo, S. Toshinori, K. Masahiro, A. New Self-Rating Scales for Depression Clinical Psychiatry, 1985; 27(6), 717–723.

24. Williams, J. B. W. A Brief Measure for Assessing Generalized Anxiety Disorder: The GAD-7. 2006 ; https://doi.org/10.1001/archinte.166.10.1092

25. Muramatsu K, Miyaoka H, Ueshima K, Muramatsu Y, Fuse K, Yoshimine H, et al. Validation and utility of a Japanese version of the GAD-7. Jpn J Psychosom Med. 2010 .1; 50.

26. Fuju, T. Tabei, Y. Shimamura, M. Yamagami, T. Factors influencing a sense of positivity towards caregiving designed exclusively for people who provide such care for elderly family members with dementia-A preliminary study to develop a scale for the assessment of a sense of positivity towards caregiving for the elderly with dementia-Bulletin of Institute for Public Welfare. 2015;12(1), 1–14

27. Lintern. T. Quality in dementia care: Evaluating Staff attitude and behavior. PHD thesis. University Of Wales Bangor; 2001

28. Mizue, S.Yutaka, M. Dawn, B. Hajime, O.Masao, K. How quality of life indices reflect the behaviors of elderly people with dementia on dementia care mapping and the relationship among well-being, ill-being and the behavior category code, Japanese Journal of Geriatrics, 2009; 49(3), 355–366

29. EuroQOL group. EuroQOL a new facility for the measurement of health-related quality of life, Health Policy, 1990; 16, 199–208

30. Ryota, I. Shinichi, N. Takamoto, U. Tetsuya, S. Taiki, S. Validity and responsiveness of the health utility measures Japanese version in health-related quality of life: Evaluation of the use of EuroQol 5-Dimension and the Health Utilities lndex Mark 3, occupational therapy, 2010; 29(6), 763–772

31. Attkisson, C. The Client Satisfaction Questionnaire (CSQ) Scales. In Measures for Clinical Practice: A Sourcebook; 1996

32. Hisateru, T. Hiroto, I. Reliability and Validity of The Japanese Version on Client Satisfaction Questionnaire, Clinical Psychiatry, 1999; 41(7), 711–717

33. Mehta KM, Gallagher-Thompson D, Varghese M, Loganathan S, Baruah U, Seeher K, et al. iSupport, an online training and support program for caregivers of people with dementia: study protocol for a randomized controlled trial in India. Trials. 2018 ;19(1):271.

34. Pinto-Bruno AC, Pot AM, Kleiboer A, Droes R-M, van Straten A. An Online Minimally Guided Intervention to Support Family and Other Unpaid Carers of People With Dementia: Protocol for a Randomized Controlled Trial. JMIR Res Protoc. 2019 ;8(10):e14106.

35. Teles S, Ferreira A, Seeher K, Freel S, Paul C. Online training and support program (iSupport) for informal dementia caregivers: protocol for an intervention study in Portugal. BMC Geriatr. 2020 ;20(1):10.

36. Xiao LD, McKechnie S, Jeffers L, De Bellis A, Beattie E, Low L-F, et al. Stakeholders’ perspectives on adapting the World Health Organization iSupport for Dementia in Australia. Dementia. 2020; 1471301220954675.

37. Oliveira D, Jacinto AF, Gratao ACM, Ottaviani AC, Ferreira CR, Monteiro DQ, et al. Translation and cultural adaptation of iSupport in Brazil. Alzheimer’s Dement. 2020;16(S7).

38. Fiordelli M, Albanese E. Preparing the ground for the adaptation of iSupport in Switzerland. Alzheimer’s Dement. 2020;16(S7):38915.

39. Baruah U, Varghese M, Loganathan S, Mehta K, Gallagher Thompson D, Zandi D, et al. A pilot randomized controlled trial on iSupport in India: Results on the use of the online program and its effectiveness. Alzheimer’s Dement. 2020;16(S7):38912.

40. Muñoz RF, Leykin Y, Barrera AZ, Brown CH, Bunge EL. The impact of phone calls on follow-up rates in an online depression prevention study. Internet Interv. 2017 June 1;8:10–4.

